# SARS-CoV-2 RNA and antibody dynamics in a Dutch household study with dense sampling frame

**DOI:** 10.1101/2021.10.06.21263384

**Authors:** Wanda G.H. Han, Arno Swart, Axel Bonacic Marinovic, Dirk Eggink, Johan Reimerink, Lisa A. Wijsman, Bas van der Veer, Sharon van den Brink, Anne-Marie van den Brandt, Sophie van Tol, Gert-Jan Godeke, Fion Brouwer, Marieke Hoogerwerf, On behalf of the Dutch FFX-COVID-19 research group, Daphne F.M. Reukers, Nynke Rots, Chantal Reusken, Adam Meijer

**Affiliations:** Centre for Infectious Disease Control, National Institute for Public Health and the Environment, Bilthoven, the Netherlands

**Keywords:** SARS-CoV-2, Household study, molecular diagnostics, serological diagnostics, infection dynamics

## Abstract

This study investigated the dynamics of SARS-CoV-2 infection and diagnostics in household members of different ages and with different symptom severity after SARS-CoV-2 exposure during the early phase of the pandemic. Households with a SARS-CoV-2 confirmed positive case and at least one child in the Netherlands were followed for 6 weeks. Naso (NP)- and oropharyngeal (OP) swabs, oral fluid and feces specimens were analyzed for SARS-CoV-2 RNA and serum for SARS-CoV-2-specific antibodies. The dynamics of the presence of viral RNA and the serological response was modeled to determine the sampling time-frame and sample type with the highest sensitivity to confirm or reject a SARS-CoV-2 diagnosis. Transmission of SARS-CoV-2 between adults and children within a household was correlated with symptom severity of index cases. In children higher viral loads compared to adults were detected at symptom onset. Early in infection, higher viral loads were detected in NP and OP specimens, while RNA in especially feces were longer detectable. SARS-CoV-2-specific antibodies have a 90% probability of detection from 7 days (total Ig) and 18 days (IgG) since symptom onset. In conclusion this study has shown that on average, children carry higher loads of virus as compared to adults early after infection. For highest probability of detection in SARS-CoV-2 diagnostics early in infection, RT-PCR on NP and OP specimens are more sensitive than on oral fluid and feces. For SARS-CoV-2 diagnostics late after infection, RT-PCR on feces specimens and serology are more valuable.

## Introduction

Severe acute respiratory syndrome coronavirus 2 (SARS-CoV-2) has spread rapidly across the world since January 2020 [1]. In the Netherlands, the first COVID-19 (the syndrome caused by SARS-CoV-2) case was detected on 27 February 2020. From March until May 2020, the Dutch government mandated a partial lockdown. This included social distancing, self-quarantine and self-isolation orders, closing of schools, bars and restaurants, and urging people to work from home [2]. Yet, households are close-contact settings with high probability of (pre/a-symptomatic) transmission of SARS-CoV-2 after introduction of the virus. In this period, a prospective cohort study was performed including 55 complete households with a RT-PCR-confirmed SARS-CoV-2 positive case (index case) and at least one child below 18 years of age. All household contacts were tested as soon as possible after a SARS-CoV-2 infection in the household was identified. At multiple timepoints, various clinical samples were collected for molecular and serological diagnostics. Using a dense sampling strategy, SARS-CoV-2 transmission and kinetics of diagnostic parameters could be closely monitored within the households. Earlier we described that the estimated Secondary Attack Rate (SAR) in this cohort that was high (35% in children, 51% in adults), with reduced susceptibility of children compared to adolescents and adults (0.67; 95%CI: 0.40-1.1) [3]. Here we looked further into the kinetics of infection.

In the present study, we use the results of the dense sampling and various molecular and serological assays to identify participants with an acute or recent SARS-CoV-2 infection to analyse household transmission patterns in relation to disease severity. Secondly, we describe the dynamics of the infection per individual based on viral RNA and antibody presence. Lastly, we compared the dynamics of the different diagnostic methods (test and sample type), by modeling the outcomes per assay in relation to the days post symptom onset (dps), disease severity and age.

## Methods

### Study protocol

A prospective cohort study was performed following households where one symptomatic household member was tested RT-PCR positive for SARS-CoV-2 in the period 24 March – 6 April 2020 [3]. In brief, persons 18 years and older testing positive for SARS-CoV-2 RT-PCR (i.e. the index case) who had at least one child in their household below the age of 18 could be included in this study (METC nr: NL13529.041.06). Table 1 describes the sampling scheme (See Reukers et al [3] for more details). We defined adults as individuals of 18 years of age or older and individuals as SARS-CoV-2 infection positive when they tested positive in at least one RT-PCR or serological assay.

**Table 1.**
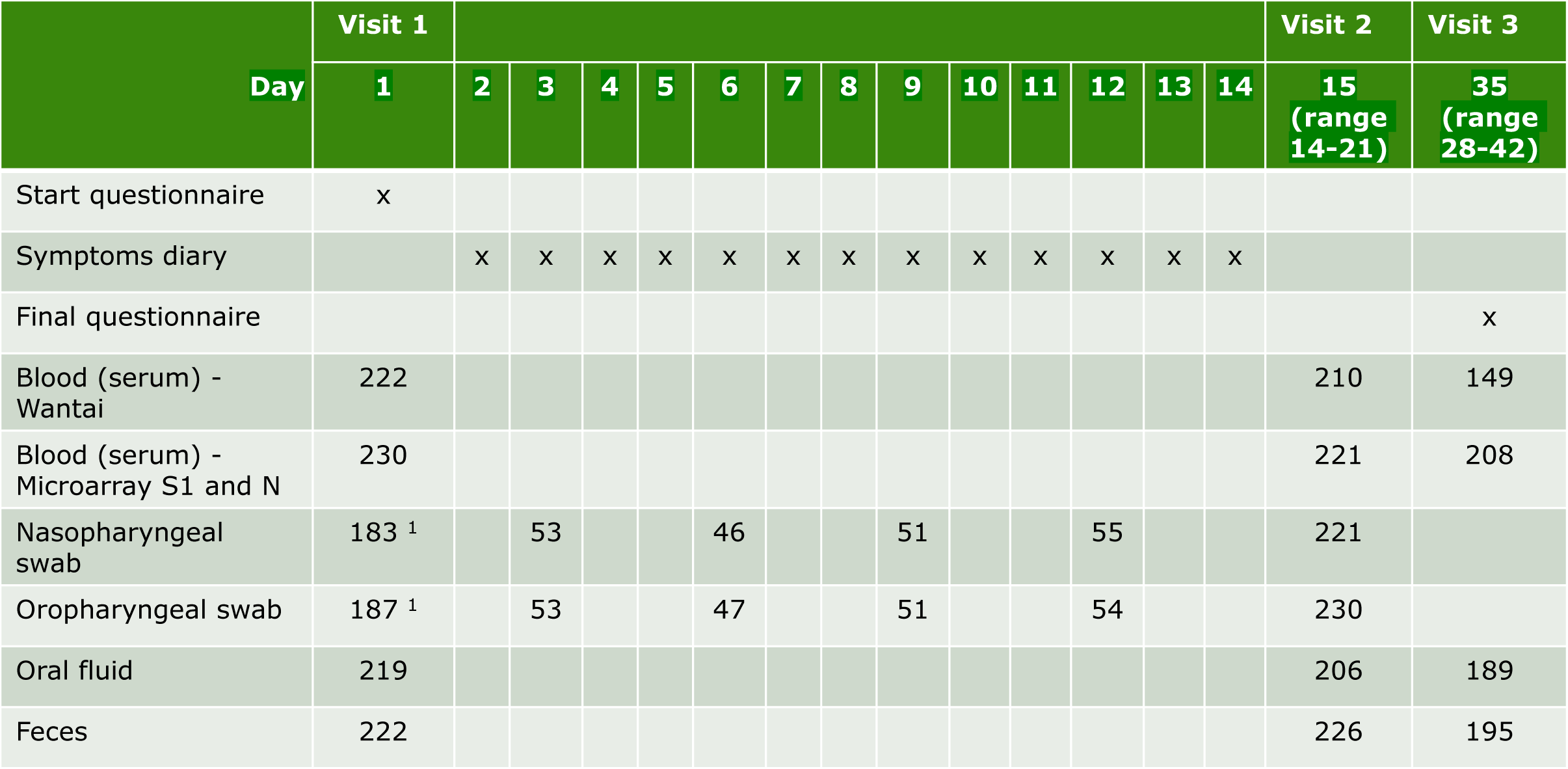
Schedule of administering questionnaires, symptom diaries and home visits for sampling. The numbers in the table indicate the amount of analyzed specimens in 242 participants. ^1^ A naso- and oropharyngeal swab was not collected for the index case at the first home visit, as these persons were already swabbed a few days before and tested SARS-CoV-2 positive.

### COVID-19 severity

The day of onset of possible COVID-19 associated symptoms, i.e. respiratory symptoms (including sore throat, cough, dyspnea or other respiratory difficulties, rhinorrhoea), fever, chills, headache, anosmia or ageusia, muscle pain, joint ache, diarrhoea, nausea, vomiting, loss of appetite or fatigue, as reported by the participant was defined 1 dps. Laboratory confirmed SARS-CoV-2 infected cases with any clinical symptoms other than pneumonia and/or requiring medical consultation were defined as mild cases. Moderate cases showed clinical signs of pneumonia, including dyspnea. Severe cases reported dyspnea and consulted a health care professional for their symptoms, or reported having been admitted to the hospital for COVID-19 [3, 4].

### Transmission categories

We categorized the household transmission patterns in three groups. In the ‘no transmission’ category, SARS-CoV-2 infection was only detected in the index case. In the ‘adult transmission’ category, SARS-CoV-2 infection was detected in adults other than the index case only. In the ‘family transmission’ category, SARS-CoV-2 infection was detected in children and possibly also adults other than the index case. In order to assess the significance of differences in severities over transmission categories a generalized linear model (GLM) for count data with Poisson family and exponential link was set up with interactions between the transmission category and severity classes.

### Molecular diagnostics

Nasopharyngeal swabs (NP) and oropharyngeal swabs (OP) were collected in gelatin-lactalbumin-yeast (GLY) viral transport medium (Mediaproducts BV, Groningen, The Netherlands), transported to the laboratory in a cooling box and stored at maximum a few days at 4° C until being processed for RT-PCR. Feces specimens were self-collected by the patient and send to the laboratory by regular mail, stored frozen at -20° C until being processed for RT-PCR. Oral fluid specimens were collected with a Oracol sponge (Malvern Medical Developments Ltd, U.K.), transported to the laboratory in a cooling box, processed for storage according to the manufacturer’s instructions, and aliquots stored frozen at -80° C until being used for RT-PCR. Total nucleic acid was extracted from NP-, OP swab, oral fluid or feces using MagNApure 96 (MP96) with total nucleic acid kit small volume (Roche). Of the feces specimens a 5% suspension was made in MEM with Hanks’ salts and penicillin and streptomycin, vortex for 15 seconds and 1 minute centrifuged at 16,000 Relative Centrifugal Force. Two-hundred µl supernatant was mixed with 275 µl MP96 lysis buffer including equine arteritis virus (EAV) internal control and yeast tRNA stabilizer. Total nucleic acid was eluted in 50 µl Tris EDTA buffer. RT-qPCR was performed on 5 µl total nucleic acid using TaqMan® Fast Virus 1-Step Master Mix (Thermo Fisher) on Roche LC480 II thermal cycler with SARS-like beta coronavirus (Sarbeco) specific E-gene primers and probe and EAV as described previously [5, 6]. As no other Sarbeco viruses are currently detected in humans, a positive Sarbeco E-gene RT-qPCR is validly taken as positive for SARS-CoV-2. For modeling purposes no detection of SARS-CoV-2 RNA was given an artificial cycle threshold (Ct) value of 40.

### Serological diagnostics

The Wantai SARS-CoV-2 total antibody ELISA (Beijing Wantai Biological Pharmacy Enterprise, Beijing, China; catalogue number WS1096) was performed according to the manufacturer’s instructions [7]. This assay is a double-antigen sandwich ELISA using the recombinant receptor-binding domain of SARS-CoV-2 as antigen. Optical density (OD) is measured at 450 nm and the antibody OD ratio for each sample is calculated as the ratio of the OD of that sample to the reading of a calibrator (included in the kit).

Sera were tested for the presence of IgG antibodies reactive with the SARS-CoV-2 S1 and SARS-CoV-2 N antigens in a protein microarray, in duplicate 2-fold serial dilutions starting at 1:20, essentially as described previously [8]. For each antigen, a 4-parameter loglogistic calibration curve was generated. Antibody titers (EC50 value) were defined as the interpolated serum dilution that gave a fluorescence intensity of 50% of the corresponding calibration curve. Raw data were processed with the R 4.04 statistical software as described previously [9].

### Modeling RT-PCR

All available RT-PCR outcomes (Table 1) were modelled by a Bayesian hierarchical model of the form

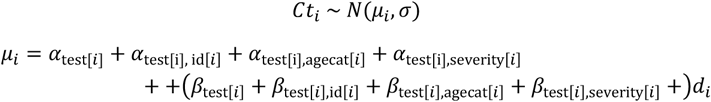

Here, *Ct*_*i*_ is the measured Ct value for sample *i*, and *σ* the overall variation. There is a part dependent on the days since onset of symptoms (*d*_*i*_) with coefficients *β*, and a constant part with coefficients *α*. Both the *α* and *β* parameters include several contributions stratified by categorical variables: id[*i*] is the person specific identifier for sample *i* enabling longitudinal modelling, agecat[*i*] is either ‘child’ or ‘adult’, severity[*i*] is either ‘asymptomatic’, ‘mild’, ‘moderate’ or ‘severe’, and test[*i*] is either ‘pcr NP, ‘pcr OP, ‘pcr oral fluid’, ‘pcr feces’, ‘wantai’, ‘microarray S1’ or ‘microarray N’.

For the id’s a hierarchical model is built (i.e. a random effect),

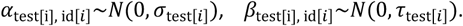

Whenever a Ct-value of 40 is encountered, we apply censoring in the model by changing the probability density function *f*(*x*; *μ*_*i*_, *σ*) into its cumulative counterpart 1 − *F*(40; *μ*_*i*_, *σ*), thereby encoding that we have an unknown Ct value which would either indicate the absence of amplifiable RNA or presence of RNA but well below the detection limit of the used RT-PCR. All parameters are given weakly informative priors, and the posterior distributions are obtained using the JAGS software [10], interfaced from R [11]. Bayesian credible intervals were obtained from the samples of the posterior as calculated by JAGS. Prediction intervals were calculated by drawing randomly from 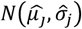, where the indicated means and standard deviations are samples from the posterior distributions. The posterior probability of being positive is modelled by 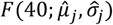. The modeling did not include the Ct values of the inclusion RT-PCR NP+OP which was performed in index cases just before start of the study, since these values are not known to us.

### Modeling serology

The dynamics of serology cannot be assumed to be linear as is the case for Ct-values. Rather, seronegative individuals have a titer (OD ratio for Wantai or EC50 for protein microarray) varying around a low value, and seropositive individuals have a titer varying around a high value. In the case of the ELISA-test and microarray-based assays used in the current study, we find that a cut-off value to distinguish seropositives and seronegatives works well, since the two components are well separated (Figure S1). Using the cut-off values 1 for Wantai (according to manufacturer’s instructions) and 10 for microarray [8], we classify each measurement *X*_*i*_ as either positive or negative. Using a Bernouilli distribution and logit link for the probability we model the outcomes as

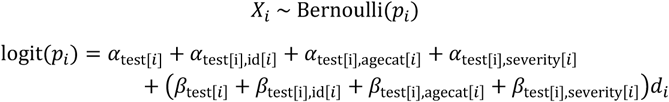

Parameter estimation proceeds analogous to the RT-PCR model.

### Assessing differences between factors

We assessed the difference between posterior estimates of parameters using the Region of Practical Equivalence (ROPE) [12-14]. The ROPE is an interval chosen based on domain knowledge that indicates values that are practically indistinguishable. For Ct-values our ROPE interval is [-1,1], which means that we consider differences between Ct-values of less than one as not meaningful. For changes in Ct-value per day (the slope) we choose [-1/7, 1/7], which means that we consider differences between Ct-values of less than one per week as not meaningful. For serology detection probability (dps) our ROPE interval is [-2,2], which means that we consider differences between days of less than 2 as not meaningful. The ROPE is compared to the 89% highest posterior density interval (HDI). When the ROPE contains the HDI, no meaningful difference exists, when the ROPE is completely outside of the HDI, there is a difference, when the ROPE and HDI overlap we withhold a decision because of too high uncertainty.

## Results

### Household transmission SARS-CoV-2

A total of 242 participants from 55 complete households were included in this study. The number of analyses performed per assay and specimen type at the various timepoints with the day of the first home visit (so the start of the study within the particular household) defined as day 1 are described in Table 1 and Table S1. To identify different transmission patterns, we visualized SARS-CoV-2 infection detection by the different assays and specimen types per participant and household in heatmaps. We identified the transmission pattern ‘no transmission’ in 16 households (Figure 1A), ‘adult transmission’ in 11 households (Figure 1B) and ‘family transmission’ in 28 households (Figure 1C). Eight of the 28 households in the ‘family transmission’ category did not show transmission to adults.

**Figure 1.**
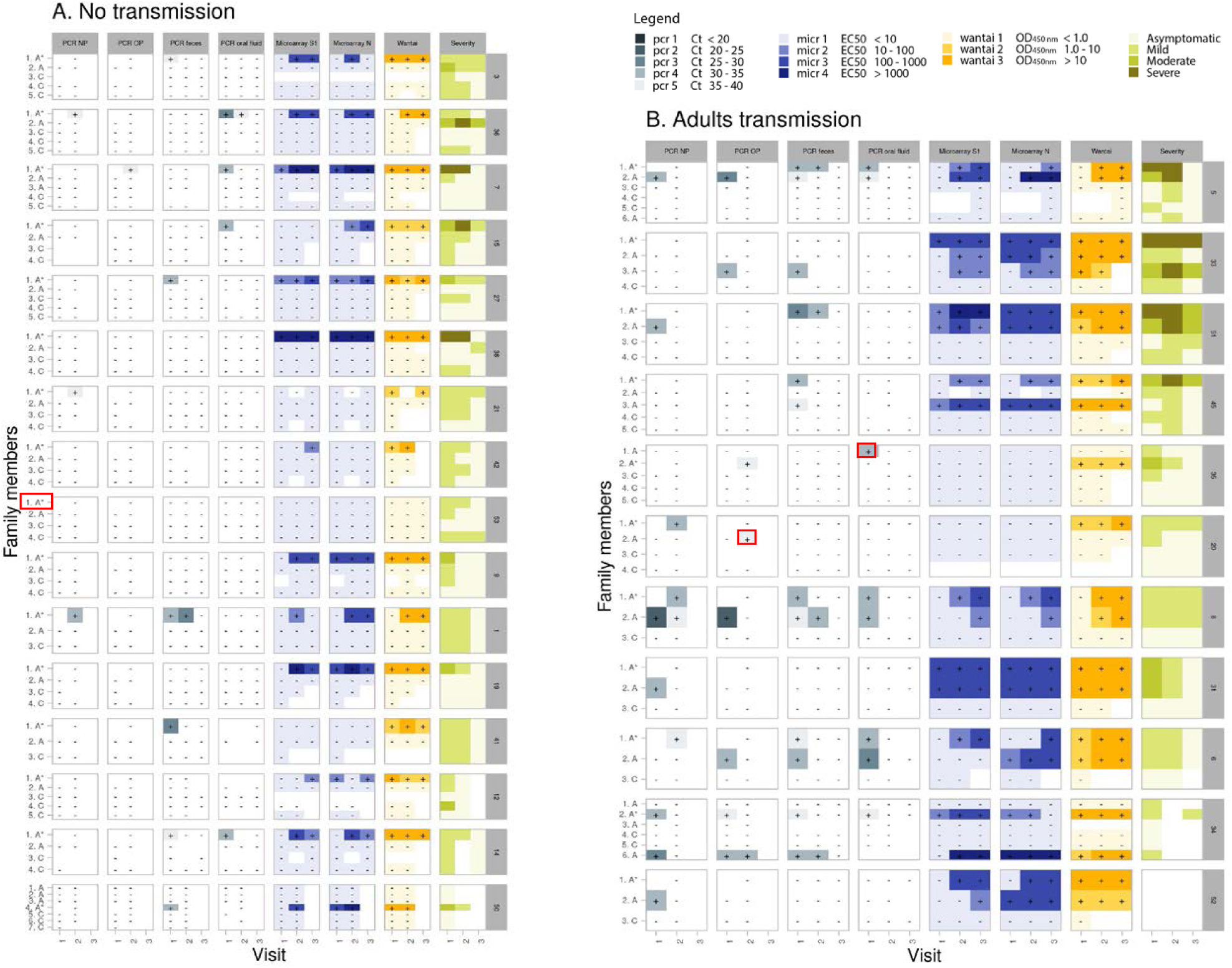
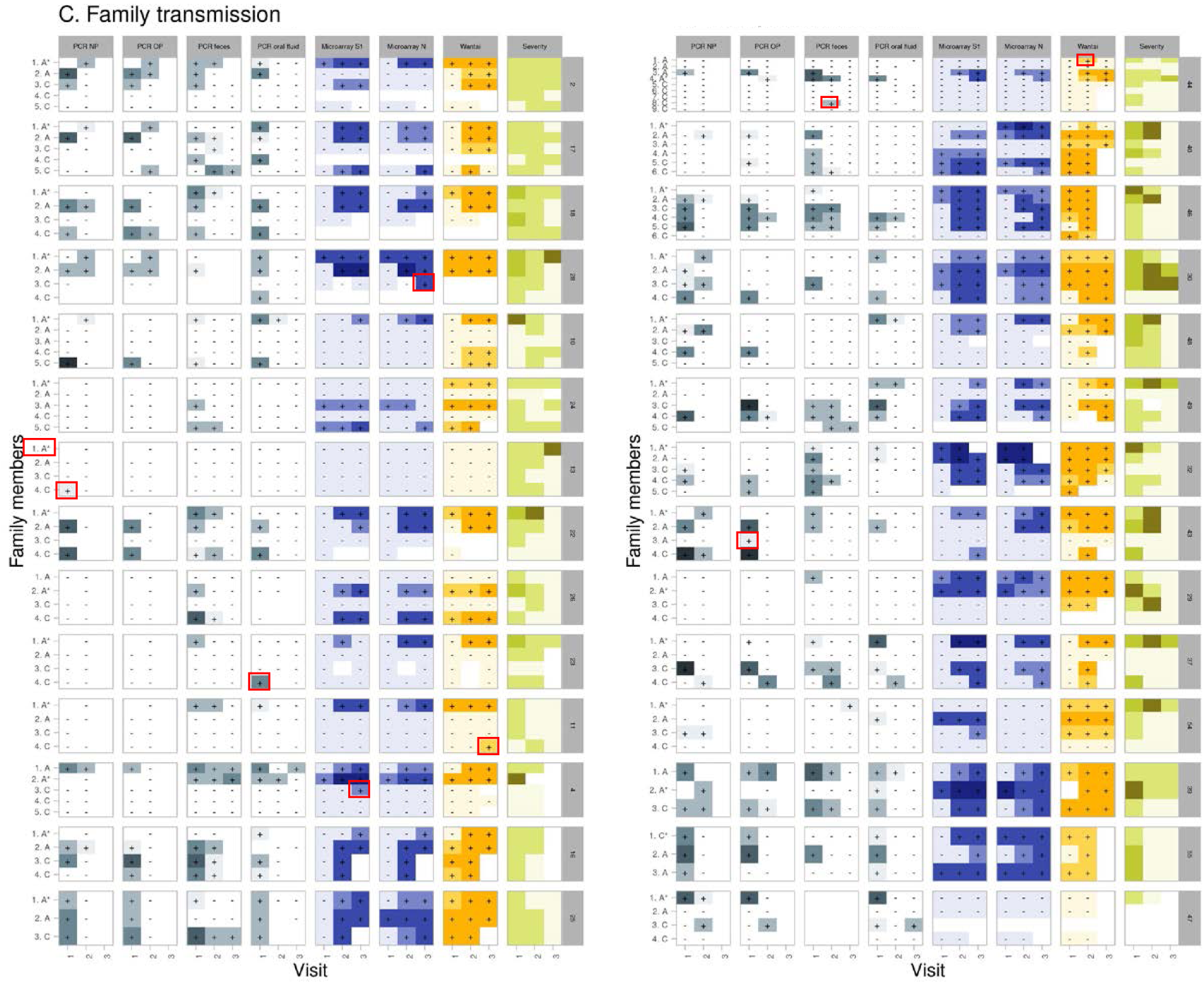
Various transmission patterns of SARS-CoV-2 infection based on different assays and specimen types collected at visits 1, 2 and 3 in households visualized in heatmaps. (A) households with no transmission. (B) household with only transmission in adults. (C) heatmaps of household with transmission in children and possibly also adults. Symptoms can be unrelated to a SARS-CoV-2 infection. On the left side, age category (A = adult and C = Child) of the participant is indicated. * Index case. Blanks = not available/tested. Red rectangle: individual with only one test positive on one timepoint. On the right side the Household ID (number) is indicated.

Symptom severity of COVID-19 index cases correlated with transmission of SARS-CoV-2 between adults and children within a household as reflected by the overrepresentation of index cases with severe symptoms in the family transmission group (p=0.03, 54% of indexes with severe COVID-19) compared to the other groups (Figure 2). In the no transmission category, more than half (56.3%) of the index cases had mild symptoms, whilst only 19% had severe symptoms. In the adult transmission category, there were 4 severe index cases out of 10 (36%).

**Figure 2.**
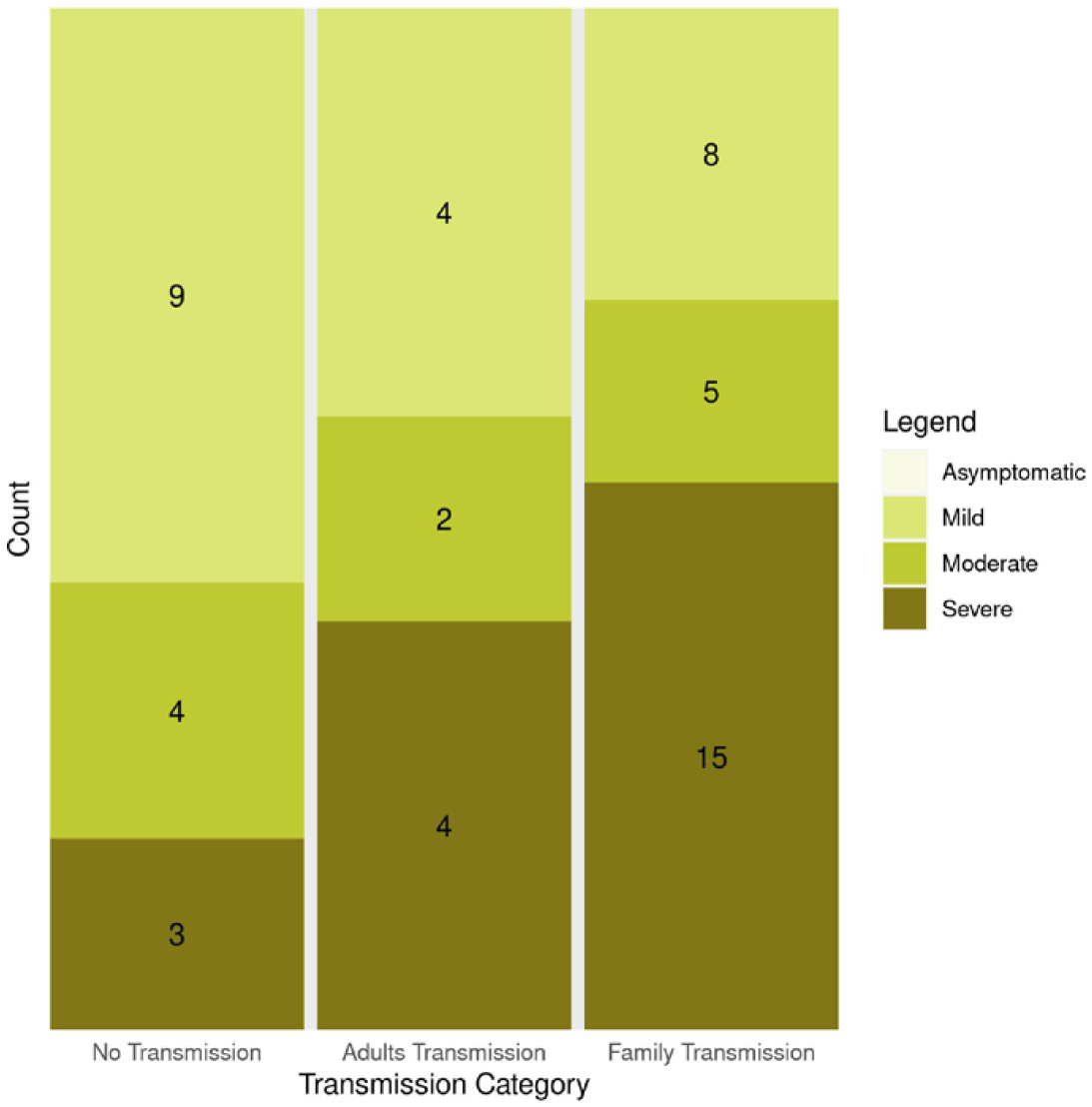
Symptom severity of COVID-19 index cases in the households. The GLM (generalized linear model) revealed that there were less individuals in the “Severe” category than in the “Mild” category (p=0.1), and that households in “Family transmission” category was overrepresented in the “Severe” category (p=0.03). Maximum severity score of index is used.

### SARS-CoV-2 infection dynamics

We investigated the SARS-CoV-2 infection dynamics in the participants of the study. Using an ‘upset plot’ [15], Figure 3 shows patterns of positive and negative results in the various molecular and serological assays. Of the 242 participants, 136 individuals were positive for SARS-CoV-2 infection by either molecular and/or serological diagnostics. Of these 136 SARS-CoV-2 infection positive individuals, 125 (91.9%) were symptomatic and severe symptomatic individuals were mainly (29 out of 32) RT-PCR- and serology-positive (Figure 3B). Most individuals were found SARS-CoV-2 positive by multiple diagnostic assays and/or materials, but 19 individuals tested positive with only one assay type and/or material (only Wantai n=5, RT-PCR NP n=3, RT-PCR oral fluid n=3, RT-PCR OP n=2, RT-PCR feces n=2, Inclusion PCR NP+OP n=2, MA-S n=1, MA-N n=1) during the study period (Figure 3A). In most of the infected cases (81.6%, 111 of the 136) both SARS-CoV-2 RNA and SARS-CoV-2-specific antibodies were detected (Figure 3B). For 12 individuals only at one timepoint during the study one positive test was found (Figure 1, red rectangles).

**Figure 3.**
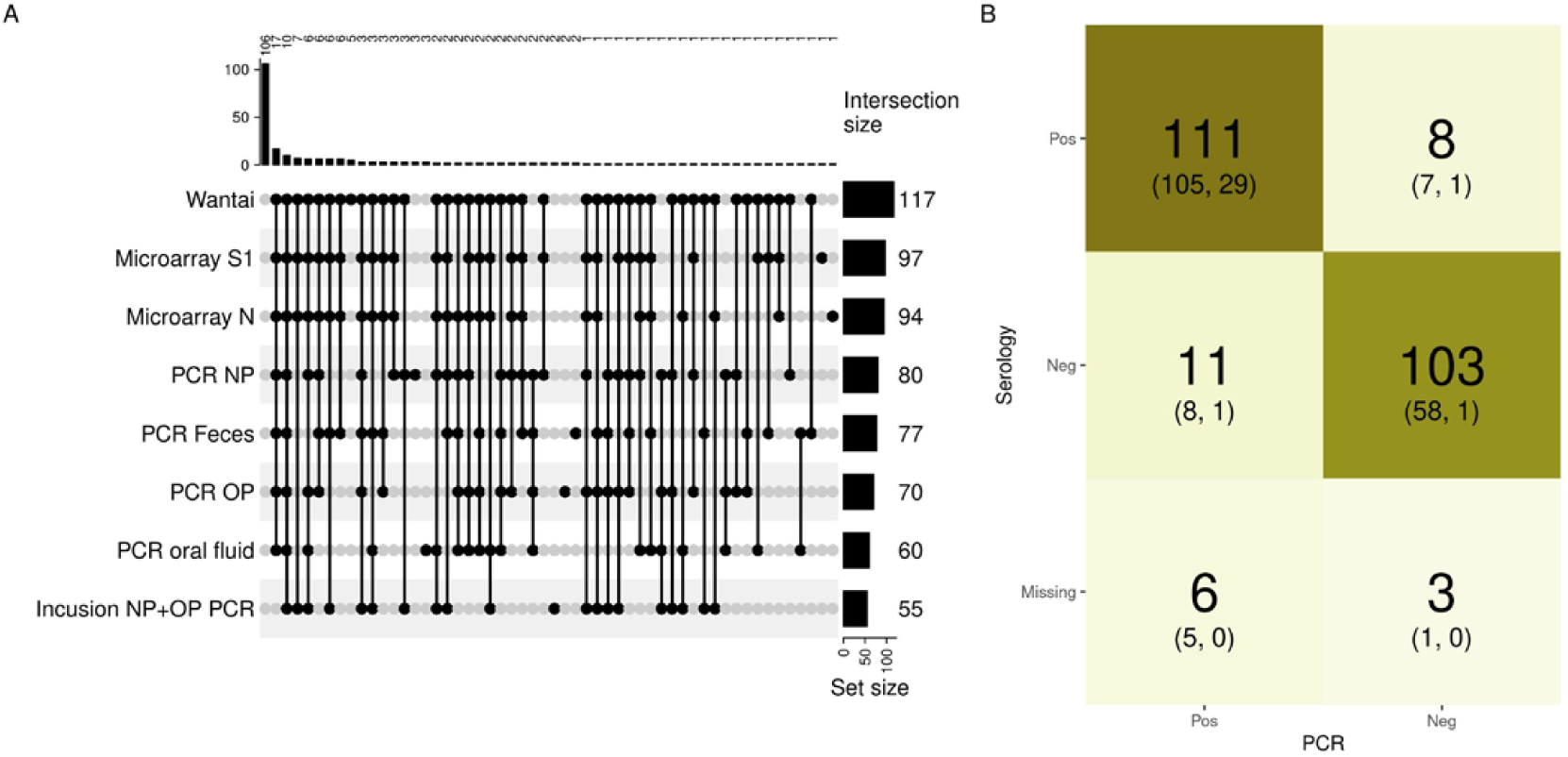
(A) Overview of the (combination of) various positive SARS-CoV-2 infection diagnosis in the 242 participants of the study cohort for all various diagnostic assays or specimens. All available RT-PCR and serology outcomes (Table 1) were incuded in these analyses.The black dots indicate a positive test at any (or multiple) moment(s) during the study, except for ‘Inclusion PCR NP+OP’ which was performed in index cases just before start of the study. The number of individuals with a particular combination of positive tests are indicated in the top of the figure. The numbers at the right after each test indicates the overall number of positive tests. (B) The number of individuals with a positive or negative tests or missing data for serological and molecular (PCR) diagnostics combined. In brackets the number of symptomatic or severe symptomatic individuals, respectively, are indicated. Color intensity is related to the frequency.

Next, for the individuals with at least one RT-PCR and one serological result at visit 1, 2 and 3 (n=198), we could analyze rough dynamics of the infection process (Figure 4). The median dps relative to visit 1 is indicated in Figure 4. Six common patterns in 173 individuals, ranked A – F based on frequency, could be identified. Within laboratory confirmed SARS-CoV-2 infected individuals, the common patterns B (n=28) and C (n=27) included individuals with a positive PCR and serological assay at visit 1. As can be expected, SARS-CoV-2 viral RNA was not detected anymore in these cases at the end of the study (4-6 weeks after inclusion), while SARS-CoV-2 antibodies remained present. Individuals with pattern E (n=12) did not have detectable SARS-CoV-2 RNA at any visit, but did have detectable SARS-CoV-2 antibodies at visit 1, 2 and 3. This pattern is in line with an earlier onset of symptoms compared to the individuals with pattern B and C, thus these individuals were included in the study later in their infection process resulting in already diminished viral RNA and present antibodies at visit 1. Pattern D (n=18) and F (n=9) included individuals with a positive RT-PCR at visit 1 and developed antibodies after visit 1. Compared to pattern B and C, these individuals reported their onset of symptoms 2-4 days later, thus at study inclusion (visit 1) they were earlier in their infection process. Pattern A included individuals with negative RT-PCR and serology results at all visits. These rough patterns underline that there are optimal time windows in which detection of SARS-CoV-2 viral RNA or SARS-CoV-2-specific antibodies are most appropriate in diagnostics. To further investigate this we used a modelling approach.

**Figure 4.**
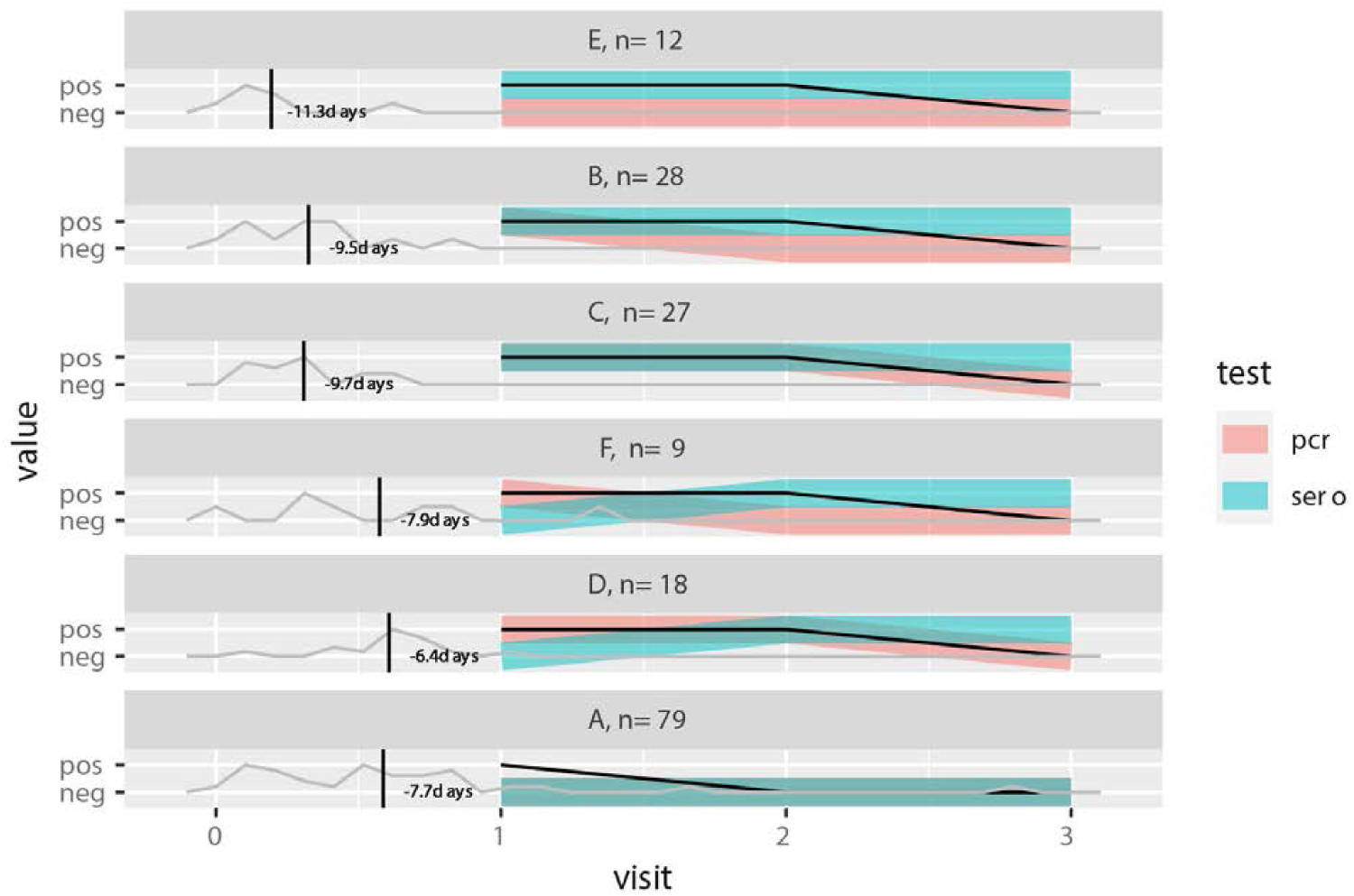
Common SARS-CoV-2 infection dynamics patterns, based on the presence or absence of a positive or negative RT-PCR or serological assay at visit 1, 2 and 3. Common patterns were named A – F based on frequency, with the number of individuals (n) displaying the pattern indicated. The median timing of onset of symptoms relative to visit 1 is indicated on the left. The black line indicates whether on average symptoms were reported at visit 1, 2 or 3.

### Dynamics SARS-CoV-2 diagnostics

Bayesian modeling on all available RT-PCR data demonstrated a difference in Ct-values at symptoms onset between adults and children (Figure 5A, B and S2-G1) and in Ct-values at symptom onset and in Ct-value increase per day between different specimen types (Figure 5C and S2-A to F). The predicted Ct value (inversely correlated with viral load), was on average 2.6 Ct lower in children (Ct 27.5; all specimens) compared to adults (Ct 30.1; all specimens) at the day of symptom onset (intercept) (Figure 5A, B and S2-G1). The decay in time in viral load (slope) was comparable between adults and children (Figure 5B and S2-G2). In line with this, there is a longer probability of SARS-CoV-2 RNA detection with increasing dps in children (99% detection until 13 dps), compared to adults (99% detection until 7.6 dps) (Figure S3A). When analyzing all ages, the predicted viral load seems slightly higher in NP and OP swabs (Ct 28.8 and 28.7) compared to feces (Ct 30.1) and oral fluid (Ct 30.7) at the day of symptom onset (intercept) (Figure 5C) indicating higher sensitivity of SARS-CoV-2 RNA detection in NP and OP specimens compared to oral fluid (Figure S2-B1 and D1) and possibly feces (Figure S2-C1 and E1). In contrast, there seems te be a slower decay (slope) in viral load in oral fluid and feces specimens (0.25 and 0.22 Ct per day) compared to NP and OP specimens (0.35 and 0.36 Ct per day) (Figure 5C). The relevance of these findings is uncertain as there is partial overlap between the ROPE and HDI (Figure S2-B1 to E1 and B2 to E2). Overall, as a most likely estimate, the estimated viral load is higher in NP and OP swabs compared to oral fuid until 21.1 dps and 19.0 dps, respectively (Figure S4-B and D), and compared to feces until 10.2 dps and 9.7 dps, respectively (Figure S4-C and E). Furthermore, there is a longer probability of SARS-CoV-2 RNA detection with increasing dps in feces (90% detection until 27 dps), compared to NP, OP and oral fluid specimens (90% detection until 19.4, 20.3 and 22.7 dps, respectively) (Figure 5D and Figure S5-C, E and F). Similar trends are shown for 50% and 10% detection probability (Figure S5 and S6). We could not find a clear correlation in severity of symptoms and the dynamics of the SARS-CoV-2 RNA detection (data not shown).

**Figure 5.**
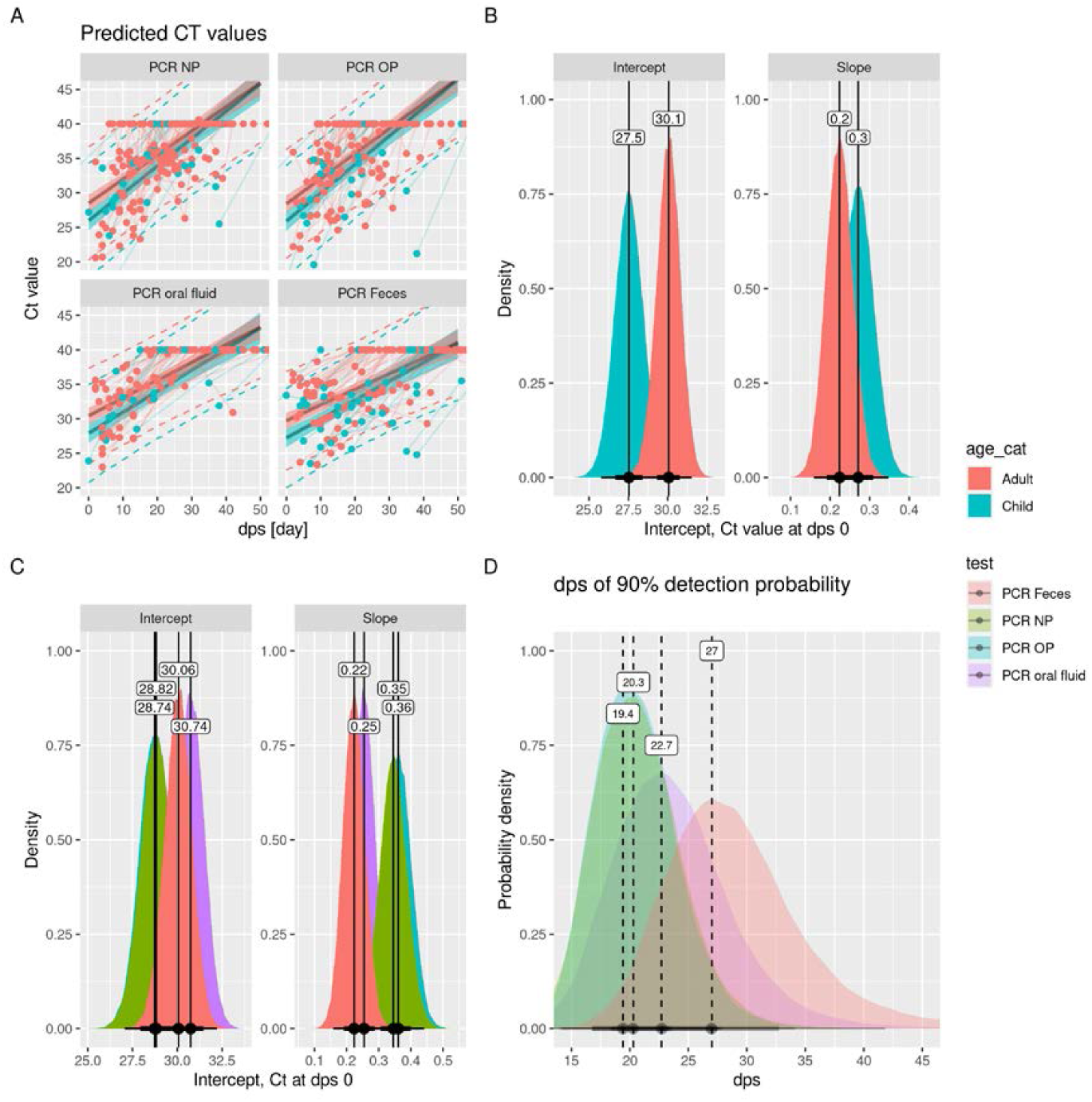
Dynamics SARS-CoV-2 infection diagnosis by RT-PCR in various specimens since symptoms onset (dps). All available RT-PCR outcomes (Table 1) were incuded in these analyses. A) Predicted viral load (Ct values RT-PCR) in relation to dps and specimen type. The shadow indicates the 95% Bayesian confidence interval and the dotted lines indicate the prediction interval (variation over individuals). B and C) Ct-value distribution at day symptom onset (intercept) and increase of Ct-value per day (slope) in relation to age category (B) and different specimens (C). D) Average dps until when different specimens have at least 90% detection probability.

Furthermore, we investigated the dynamics of SARS-CoV-2 antibody detection (Figure 6). The Wantai assay (total Ig) demonstrated a higher sensitivity for detection of anti-S1 antibodies than the micro-array (IgG) as the probability for detection was earlier using Wantai upon onset of illness (Figure 6A and S7-A). The dps at which 90% detection probability was reached for Wantai was 7.1 compared to 16.9 and 18 for Nucleoprotein (N)- and S1-protein microarray respectively (Figure 6B). The protein microarray for S1 and N had comparable sensitivity, in line with a previous study [8]. The probability of detecting N-specific IgG antibodies in children was delayed by 3.0 days (at 90% probability detection) versus adults (Figure 6C and D), while this was not the case for detection of S1-specific antibodies (Figure 6C). The relevance of this finding is uncertain as there is much overlap between the distributions in children and adults (Figure S8). The N-specific IgG antibody titers (at visit 3; convalescent phase) were not significantly lower in children compared to adults (Figure S9). Furthermore, we could not find a correlation in severity of symptoms and the dynamics of the SARS-CoV-2 infection detection by the serological assays (data not shown).

**Figure 6.**
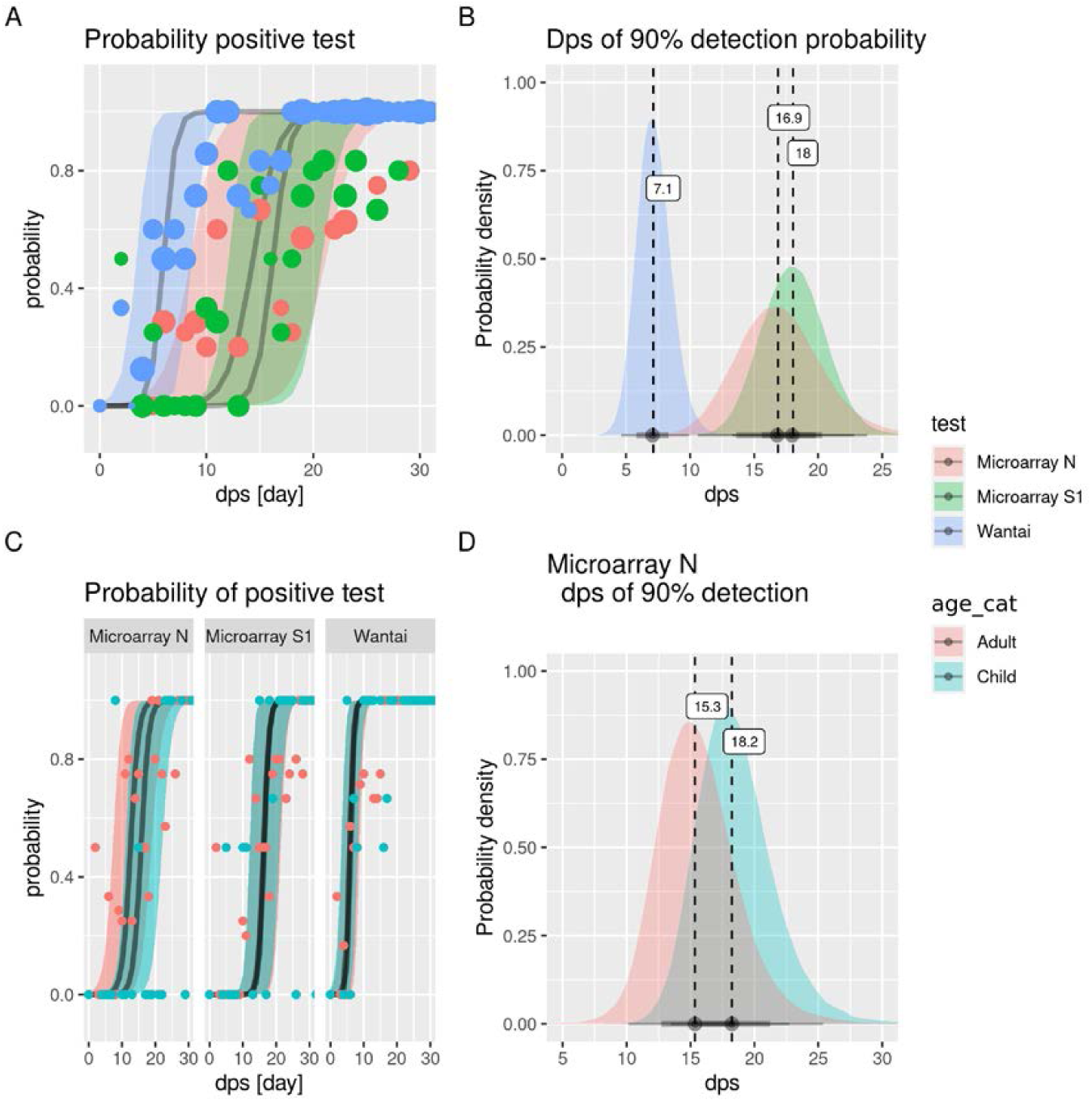
Dynamics SARS-CoV-2 infection diagnosis by different serological assays. All available serology outcomes (Table 1) were incuded in these analyses. Probability SARS-CoV-2-specific antibody detection (A) and average dps from when the Wantai (Spike-specific IgM and IgG), microarray S1 (Spike-specific IgG) and microarray N (Nucleoprotein-specific IgG) assays have at least 90% detection probability (B) for all ages. C) Probability SARS-CoV-2-specific antibody detection by Wantai, microarray S1 and microarray N in adults and children D) Average dps from when microarray N has at least 90% detection probability for adults and children. The shadows in (C) and (D) indicate the 95% Bayesian confidence interval.

## Discussion

We studied SARS-CoV-2 RNA and antibody kinetics in a household cohort during the early phase of the pandemic using an unusual dense sampling schedule allowing for high resolution analysis. For highest probability of detection of SARS-CoV-2 RNA by RT-PCR early in infection NP and OP are more suitable than oral fluid and feces. SARS-CoV-2-specific antibodies have a 90% probability of detection from 7 dps with the Wantai assay and 18 dps with the microarray S1 and N assay. This study has been performed in a naïve population during the early phase of the pandemic. Although SARS-CoV-2 vaccination, previous infection and the circulation of other SARS-CoV-2 variants may influence the dynamics of SARS-CoV-2 infection and thereby diagnostics, our study provides valuable reference insights into this subject.

Households present close-contact settings with high risk of SARS-CoV-2 transmission after introduction of SARS-CoV-2 in the household [3, 16]. We observed a positive correlation between transmission of SARS-CoV-2 between adults and children and the severity of disease in the household indexes. This is in line with studies that report that the severity of SARS-CoV-2 infection of the index case was associated with higher infectiousness [16, 17]. It should be noted that at the time of the study, SARS-CoV-2 testing in the Netherlands was limited to symptomatic healthcare workers and symptomatic vulnerable individuals. The index cases were thus mainly symptomatic healthcare workers [3]. As schools and daycare centres were closed during the study period, transmission outside the households among children was minimalized. Due to limited sample size, we could only categorized children as those of 17 years of age or younger for the Bayesian hierarchical modelling. It would be of interest to stratify the children in more groups, e.g. primary school age and adolescent age.

Seroconversion rates in mild to severe symptomatic SARS-CoV-2 RT-PCR positive cases have been reported in the range of 93-100% after 3-4 week [18-20]. During the course of this study, a vast majority of RT-PCR confirmed SARS-CoV-2 infected participants (91.0%, 111/122) developed SARS-CoV-2 specific antibodies (Figure 3B). In 6 RT-PCR positive cases, serology data was missing. In 11 of 122 (9.0%) RT-PCR confirmed SARS-CoV-2 infected individuals there was no seroconversion (Figure 3B). These individuals may have experienced a relative mild infection, or in the cases with only one positive RT-PCR test (including the available results of the extra sampling between visit 1 and 2, n=8) and no evidence of antibody response, the SARS-CoV-2 diagnosis of these individuals is disputable. Four of the 8 RT-PCR-negative cases that did have SARS-CoV-2-specific antibodies during the study period, showed only one positive serological test, suggesting also a disputable SARS-CoV-2 diagnosis. In the other 4 individuals, SARS-CoV-2-specific antibodies were detected at multiple visits including at visit 1, therefore likely having experienced infection before inclusion. Nevertheless, technically there was no evidence of false positivity as all negative controls had correct results. Therefore, the disputable results remain unexplained. Transient exposure without established infection and rapid waning of immune response might be one explanantion.The individuals with negative RT-PCR and serology results at all visits were probably not infected with SARS-CoV-2 (Figure 3B and Figure 4, pattern A). Those reporting symptoms were not tested for alternative diagnoses (i.e. other respiratory viruses). From national surveillance reports we known that also other viruses causing COVID-19-like acute respiratory infection symptoms circulated (https://www.rivm.nl/virologische-weekstaten). Especially in March 2020, the first study month, before the COVID-19 measures were put in place. Later on rhinoviruses continued circulating during the measures.

In the current study we used Bayesian hierarchical models to determine the sampling time-frame and sample type with the highest sensitivity to confirm or reject a SARS-CoV-2 diagnosis. Due to the close-contact settings with high probability of (pre/a-symptomatic) transmission of SARS-CoV-2 after introduction of the virus in households and the longitudinal dense sampling performed in this study, the analysis yielded results with relatively narrow credibility intervals which support our findings. For highest probability of a SARS-CoV-2 diagnosis (lower Ct), RT-PCR on NP and OP specimens were more suitable than feces and oral fluid until 10 dps and 20 dps, respectively (Figure S4C, E and B, D). Our study confirms that NP and OP (or combined NP+OP) were the preferred sample type for RT-PCR-based SARS-CoV-2 diagnostics within 1 week upon onset of symptoms [21-23]. A systematic review concluded that of the alternative specimens to NP and OP swabs, oral fluid (saliva) has an estimated sensitivity of 83.9% (95% CI: 77.4-88.8) and specificity of 96.4% (95% CI: 89.5-98.8) compared to reference NP and OP swabs in nucleic acid assays [23]. The sensitivity and specificity of feces specimens seems much lower, although limited data is available [23]. For SARS-CoV-2 diagnostics late in infection or in past infections, RT-PCR on feces and oral fluid specimens are more valuable than NP and OP specimens, since the presence of viral RNA in especially feces remain present over a longer time compared to NP and OP swab specimens. This is in line with findings of other studies that indicate that SARS-CoV-2 RNA can be detected up to 126 days in feces compared to 83 days in respiratory specimens and that beyond 10 dps, feces sampling may be preferred [22, 24, 25]. Although SARS-CoV-2 RNA can remain present in respiratory and feces specimens for a long time, the duration of presence of viable virus is relatively short-lived [26, 27]. Alternatively, from a week after symptom onset testing for the presence of SARS-CoV-2 Spike-specific total Ig using Wantai ELISA can confirm a recent or past SARS-CoV-2 infection. SARS-CoV-2 diagnostics using protein microarray detecting SARS-CoV-2 Spike- and Nucleoprotein-specific IgG antibodies, is useful 2 weeks after infection or symptom onset.

The infection dynamics of SARS-CoV-2 may be influenced by characteristics of the tested population, such as age and the severity of COVID-19. We, however, could not find a clear correlation in severity of symptoms or age and the RNA and SARS-CoV-2-specific antibody kinetics, although our study may be underpowered to detect these differences (data not shown). Children in general report milder symptoms compared to adults (Table S1) [3]. It is known that with age, the expression of ACE-2 increases in nasal epithelium [28]. Since SARS-CoV-2 uses the ACE-2 receptor for host entry, a lower expression of *ACE-2* in children relative to adults might explain the lower susceptibility and milder infection course in children. However, our findings and other studies show that viral loads in children are similar or higher than viral loads in adults [29, 30]. In our study children displayed lower Ct values (higher viral loads) at the day of symptom onset compared to adults, while the decay in viral load was comparable (Figure 5). This suggests that if children become infected with SARS-CoV-2, they can carry high loads of virus for a longer time compared to adults. Therefore, children are potentially longer infectious than adults after symptom onset. Whether this observation holds for new Variants of concern (VOC) e.a. delta, warrants further investigation. No clear differences between adults and children were found in the dynamics of SARS-CoV-2 serology, yet the detection of N-specific antibodies seems slightly delayed in children compared to adults (Figure 6C, D and S8). A study showed a reduced breadth of anti-SARS-CoV-2-specific antibodies, predominantly generating IgG antibodies specific for the S protein but not the N protein in children compared to adults [31]. Whether this has consequences for the development of immunity to SARS-CoV-2 is not yet clear.

In summary, our study allowed for a high resolution analysis of the sensitivity of molecular and serology-based detection of recent SARS-CoV-2 infections due to the unusual dense sampling strategy in a confined setting. For highest probability of SARS-CoV-2 diagnostics early in infection, PCR on NP and OP specimens are in favor over oral fluid and feces. For SARS-CoV-2 diagnostics late in infection or in past infection, RT-PCR on feces specimens and serology are more valuable. Children seem to carry higher loads of virus for a prolonged time in comparison to adults. The data presented here strengthen the evidence-basis for SARS-CoV-2 testing strategies.

## Data Availability

No external datasets were used in the manuscript

## Acknowledgements

We thank the Public Health Service Utrecht for assistance in the recruitment of households. We thank Alper Çevirgel, Anneke Westerhof, Anoek Backx, Elma Smeets-Roelofs, Elsa Porter, Elske Bijvank, Francoise van Heiningen, Gabriel Goderski, Harry van Dijken, Helma Lith, Hinke ten Hulscher, Ilse Akkerman, Ilse Schinkel, Jeroen Hoeboer, Jolanda Kool, Josine van Beek, Joyce Greeber, Kim Freriks, Lidian Izeboud, Lisa Beckers, Liza Tymchenko, Maarten Emmelot, Maarten Vos, Margriet Bisschoff, Marit de Lange, Marit Middeldorp, Marjan Bogaard, Marjan Kuijer, Martien Poelen, Nening Nanlohy, Olga de Bruin, Rogier Bodewes, Ruben Wiegmans, Sakinie Misiedjan, Saskia de Goede, Titia Kortbeek, and Yolanda van Weert for assistance in logistics and laboratory analyses. We thank Bettie Voordouw for critical review of manuscript. The Dutch FFX-COVID-19 research group of the Centre for Infectious Disease Control, National Institute for Public Health and the Environment, the Netherlands, includes the following members: Arianne B. van Gageldonk-Lafeber, Wim van der Hoek, Susan van den Hof, Adam Meijer, Daphne F.M. Reukers, Chantal Reusken, Inge Roof and Nynke Rots.

This study was funded by the Dutch Ministry of Health, Welfare, and Sport (VWS).

None of the authors has any potential conflicts of interest to disclose.

