## Supplemental Material for "SARS-CoV-2 RNA and antibody dynamics in a Dutch household study with dense sampling frame"

***Table S1***

| Positivity and severity by age class | | | |
|---|---|---|---|
| | All | Adult | Children |
| cohort | | | |
| n | 242 | 125 | 117 |
| age | 27 (SD=18) | 42 (SD=11) | 10 (SD=4) |
| gender (male) | 114 (47%) | 59 (47%) | 55 (47%) |
| Visit 1 | | | |
| Asymptomatic | 66 (27%) | 17 (14%) | 49 (42%) |
| Mild | 119 (49%) | 60 (48%) | 59 (50%) |
| Moderate | 40 (17%) | 34 (27%) | 6 (5%) |
| Severe | 11 (5%) | 11 (9%) | 0 (0%) |
| PCR NP pos | 46 (25%) | 25 (32%) | 21 (20%) |
| PCR OP pos | 45 (24%) | 24 (31%) | 21 (19%) |
| PCR Feces pos | 71 (31%) | 48 (39%) | 23 (21%) |
| PCR oral fluid pos | 59 (27%) | 41 (36%) | 18 (17%) |
| Microarray S1 pos | 29 (13%) | 25 (20%) | 4 (4%) |
| Microarray N pos | 34 (15%) | 32 (26%) | 2 (2%) |
| Wantai pos | 79 (36%) | 61 (50%) | 18 (18%) |
| Visit 2 | | | |
| Asymptomatic | 97 (40%) | 32 (26%) | 65 (56%) |
| Mild | 98 (40%) | 59 (47%) | 39 (33%) |
| Severe | 24 (10%) | 22 (18%) | 2 (2%) |
| PCR NP pos | 28 (13%) | 22 (18%) | 6 (6%) |
| PCR OP pos | 18 (8%) | 11 (9%) | 7 (7%) |
| PCR Feces pos | 35 (15%) | 15 (13%) | 20 (18%) |
| PCR oral fluid pos | 8 (4%) | 6 (6%) | 2 (2%) |
| Microarray S1 pos | 83 (38%) | 62 (50%) | 21 (22%) |
| Microarray N pos | 80 (36%) | 65 (52%) | 15 (15%) |
| Wantai pos | 113 (54%) | 84 (69%) | 29 (33%) |
| Visit 3 | | | |
| Asymptomatic | 162 (67%) | 73 (58%) | 89 (76%) |
| Mild | 29 (12%) | 21 (17%) | 8 (7%) |
| Moderate | 9 (4%) | 9 (7%) | 0 (0%) |
| Severe | 4 (2%) | 3 (2%) | 1 (1%) |
| PCR NP pos | 0 (NaN%) | 0 (NaN%) | 0 (NaN%) |
| PCR OP pos | 0 (NaN%) | 0 (NaN%) | 0 (NaN%) |
| PCR Feces pos | 6 (3%) | 3 (3%) | 3 (3%) |
| PCR oral fluid pos | 2 (1%) | 1 (1%) | 1 (1%) |
| Microarray S1 pos | 90 (43%) | 68 (57%) | 22 (25%) |
| Microarray N pos | 86 (41%) | 66 (55%) | 20 (22%) |
| Wantai pos | 88 (59%) | 75 (69%) | 13 (32%) |

Table S1. Study characteristics. Description age (mean; (sd)), gender (n male; (%)), symptom severity (n; (%)) and SARS-CoV-2 positive diagnosis per test (n; (%)) for the different age cohorts.

***Figure S1***

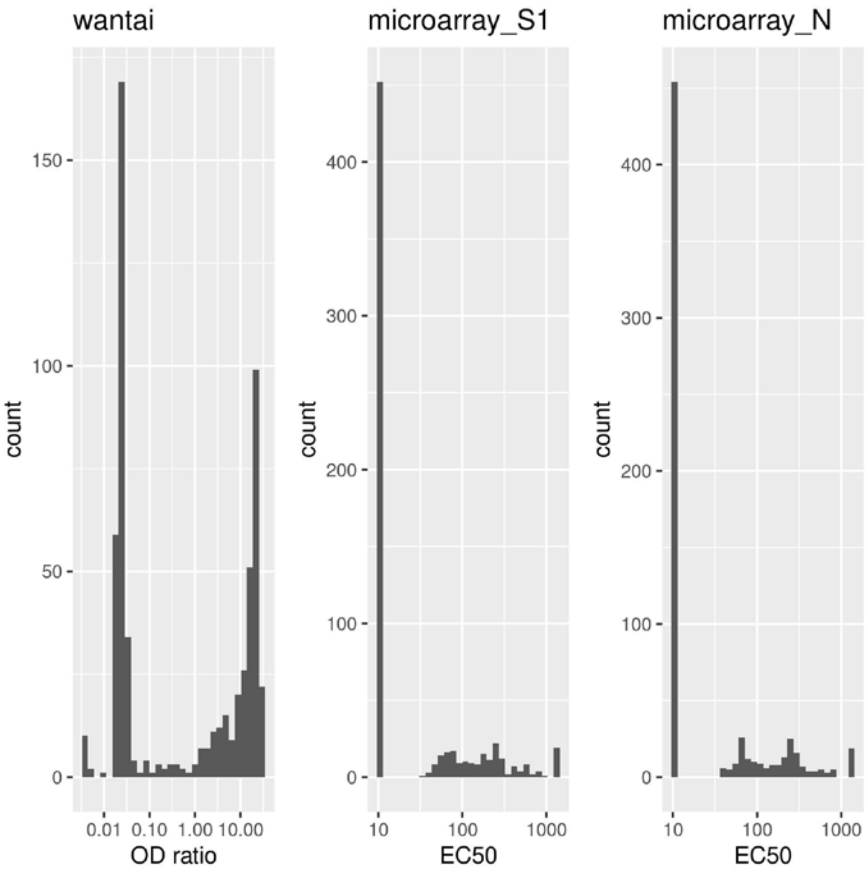


Figure S1. Histogram of Wantai OD ratios and microarray EC50 values. A clear separation between negative and positive components can be observed

*Figure S2*

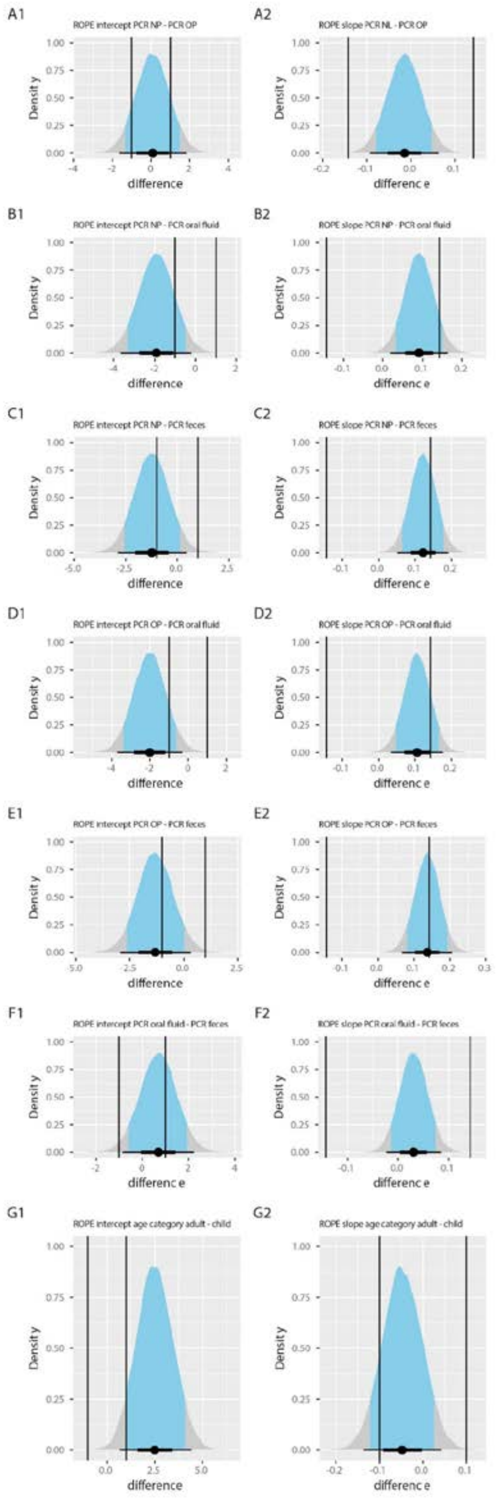


Figure S2: ROPE analyses for assessing differences between factors in Figure 5. For Ct-values our ROPE interval is [-1,1] (vertical lines, left panels A1 – G2), which means that we consider differences between Ct-values of less than one as not meaningful. For changes in Ct-value per day (the slope) we choose an interval of [-1/7, 1/7] (vertical line, right panels A2 – G2), which means that we consider differences between Ct-values of less than one per week as not meaningful. When the ROPE is outside the 89% highest posterior density interval (HDI) (blue area), there is a difference.

*Figure S3*

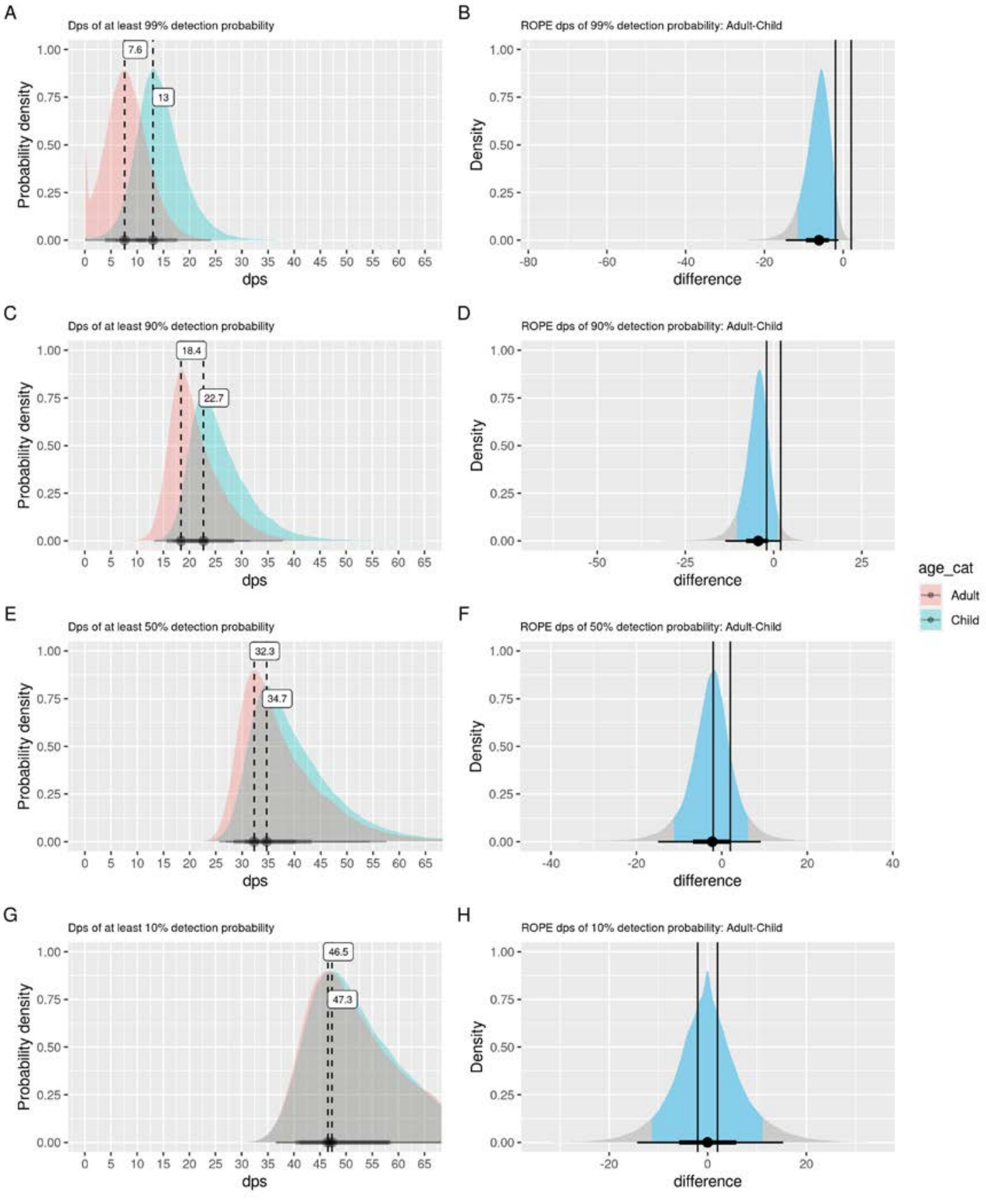


Figure S3: Dynamics SARS-CoV-2 infection diagnosis by RT-PCR in adults and children. Left panels: Average dps until when adults and children have detection probability of at least 99% (A), 90% (C), 50% (E) or 10% (G). Right panels: ROPE analyses for assessing differences between adults and children. Analyses for dps of 99% (B), 90% (D), 50% (F) and 10% (H) detection probability are shown. For Ct-values our ROPE interval is [-1,1] (vertical lines), which means that we consider differences between Ct-values of less than one as not meaningful. When the ROPE is outside the 89% highest posterior density interval (HDI) (blue area), there is a difference.

***Figure S4***

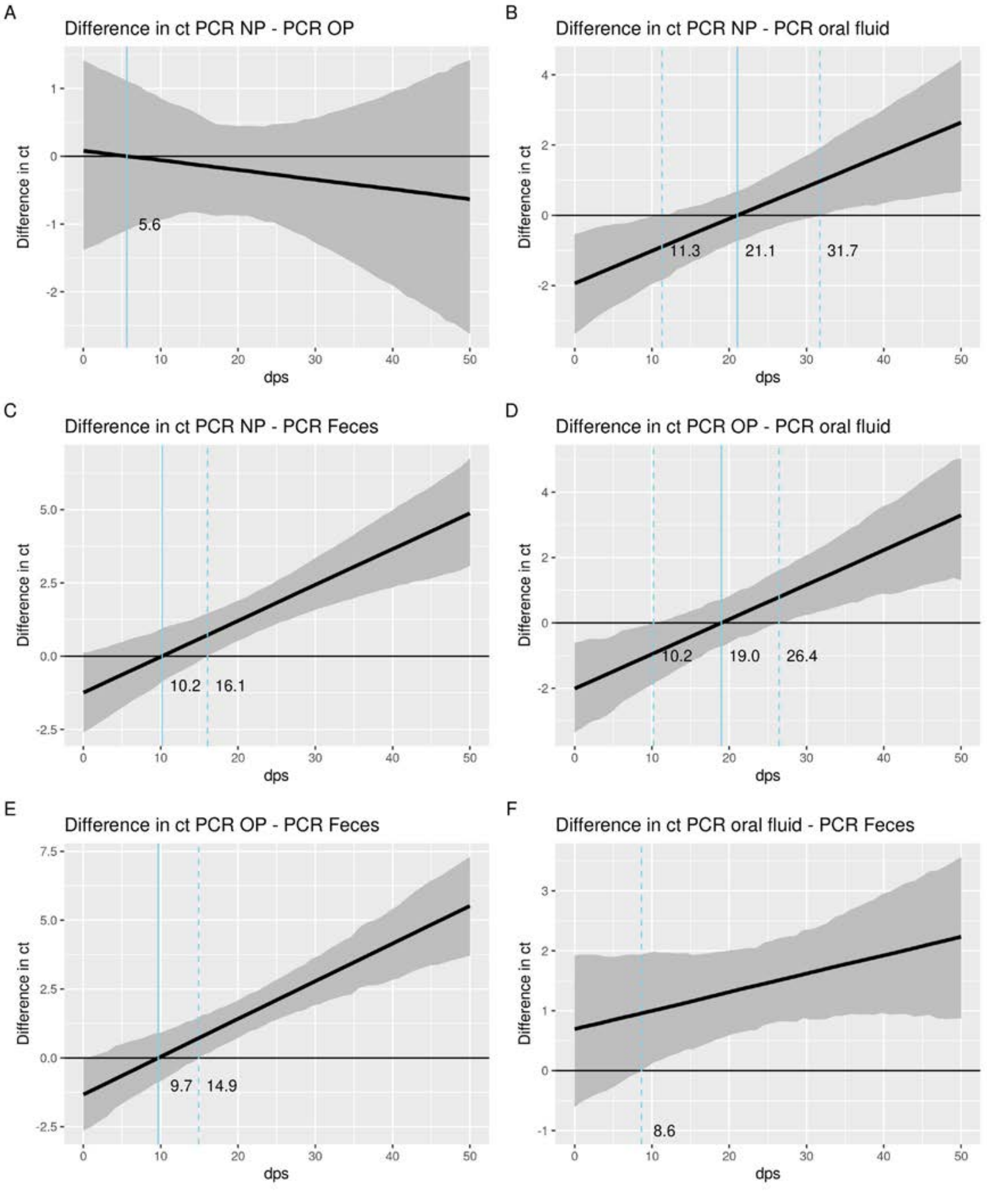


Figure S4: Differences in Ct between RT-PCR speciments in relation to dps. The black solid line indicates the median difference between the two assays (assay A – assay B) indicated, the shaded region is the 89% bayesian credible interval. For reference the line y=0 is drawn, where this line crosses the solid line of median difference, the ct values of the two assays are equal. The dps corresponding to this crossing point is indicated by a blue solid vertical line. The dashed lines indicate where y=0 crosses the top and bottom quantiles of the credible interval. To the left of the leftmost dashed line we are sure that assay A has a smaller Ct then assay B, and conversely larger to the right of the rightmost line. In between those lines we are not certain, although more area of the credible interval below zero gives more credibility to a smaller Ct for assay A.

*Figure S5*

ROPE dps of 90,50,10% detection probability.

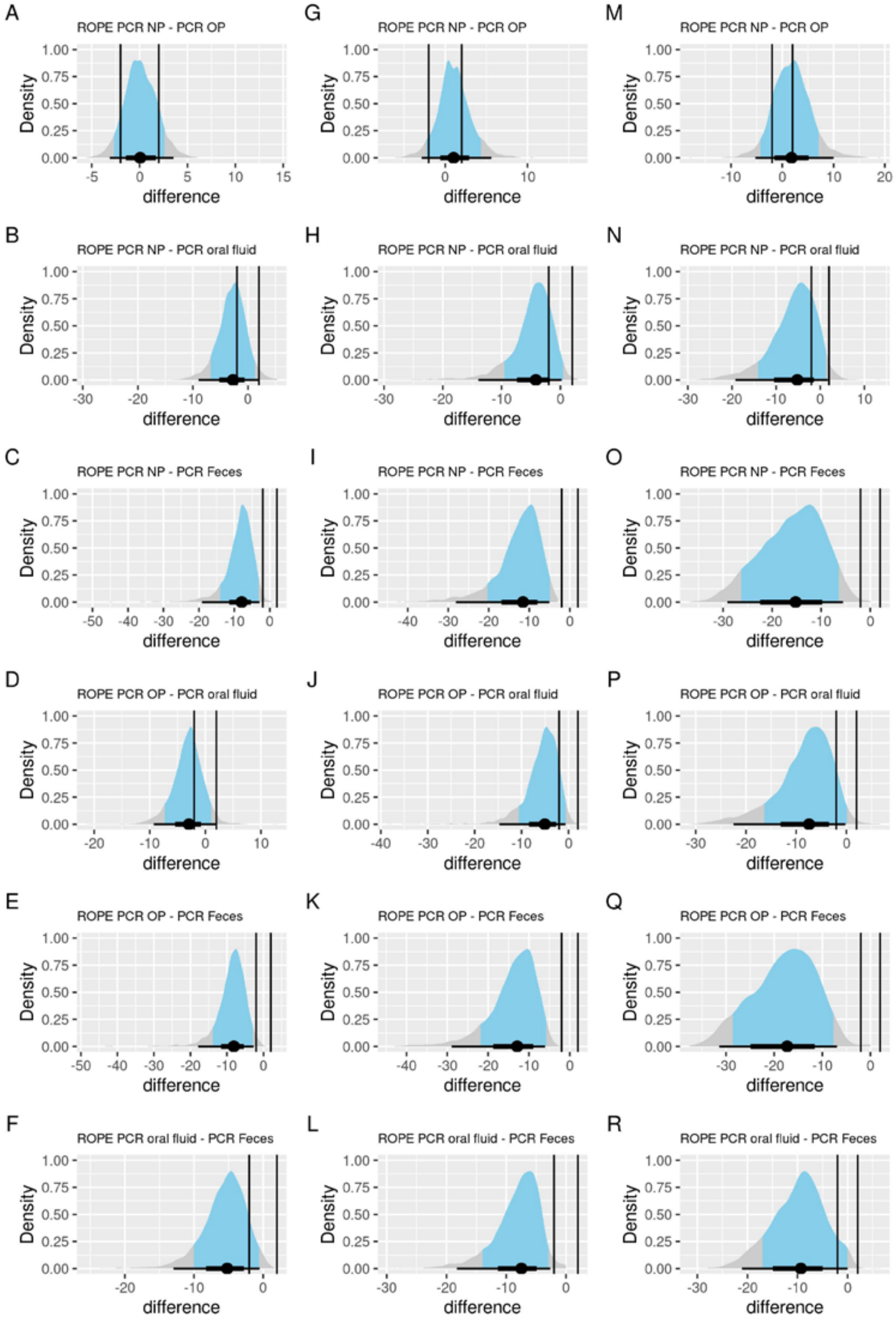


Figure S5: ROPE analyses for assessing differences between factors in Figure 5D and S4. Analyses for dps of 90% detection probability (left panels, A-F), 50% probability (middle panels, G-L) and 10% probability (right panels, M-R) are shown. For Ct-values our ROPE interval is [-1,1] (vertical lines), which means that we consider differences between Ct-values of less than one as not meaningful. When the ROPE is outside the 89% highest posterior density interval (HDI) (blue area), there is a difference.

*Figure S6*

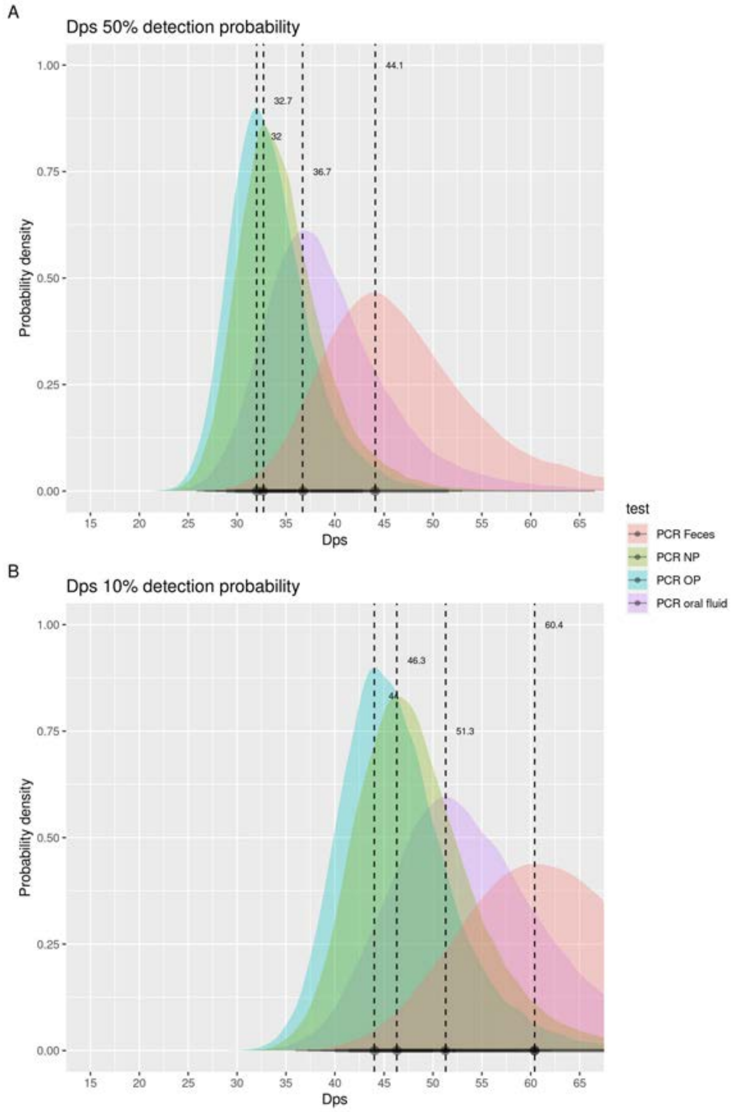


Figure S6: Dynamics SARS-CoV-2 infection diagnosis by RT-PCR in various specimens. Average dps until when different specimen have at least 50% (A) or 10% (B) detection probability.

*Figure S7*

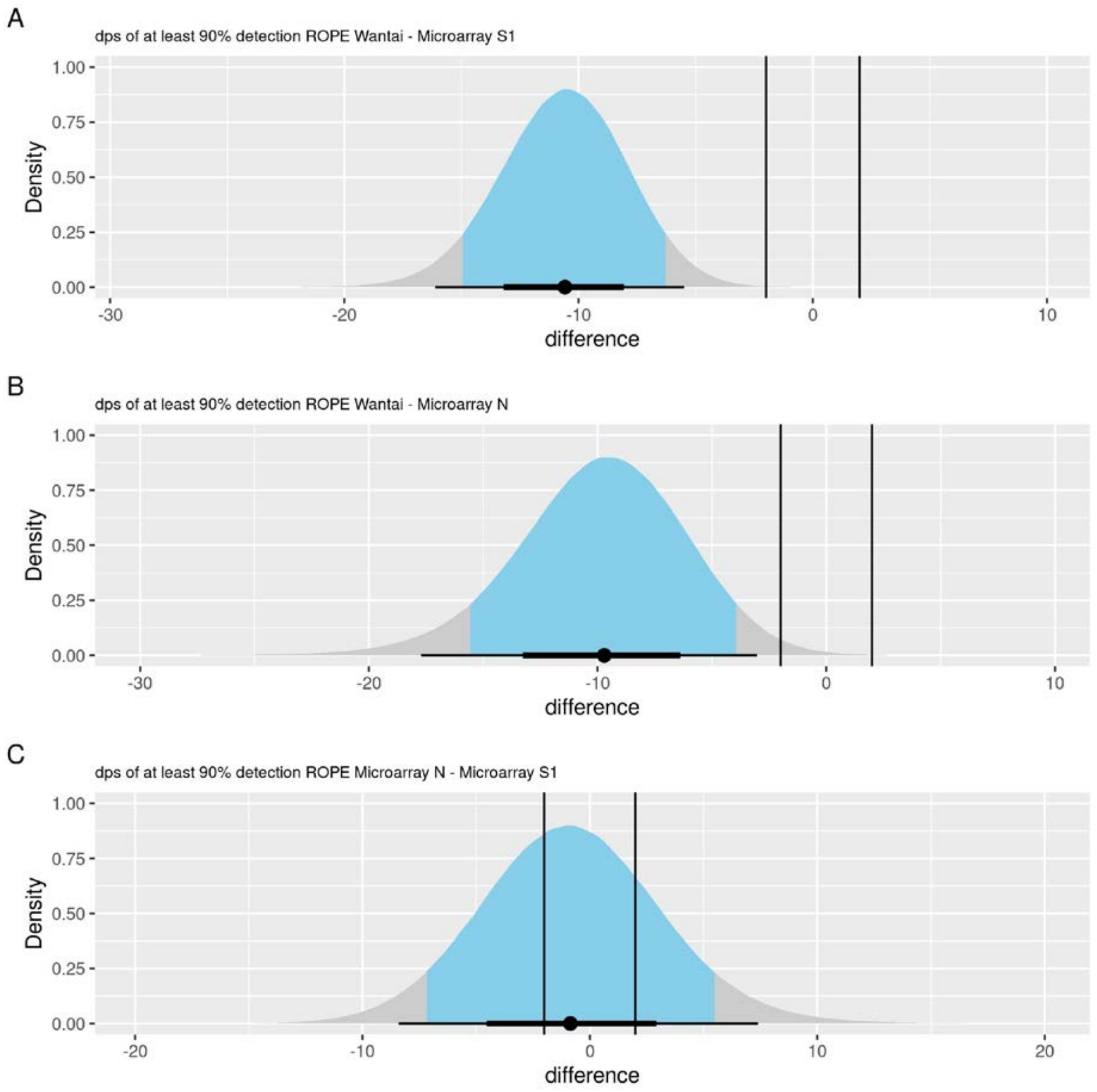


Figure S7: ROPE analyses for assessing differences between factors in Figure 6A and B. For serology detection probability (in dps) our rope interval is [-2,2], which means that we consider differences between days of less than 2 as not meaningful.

*Figure S8*

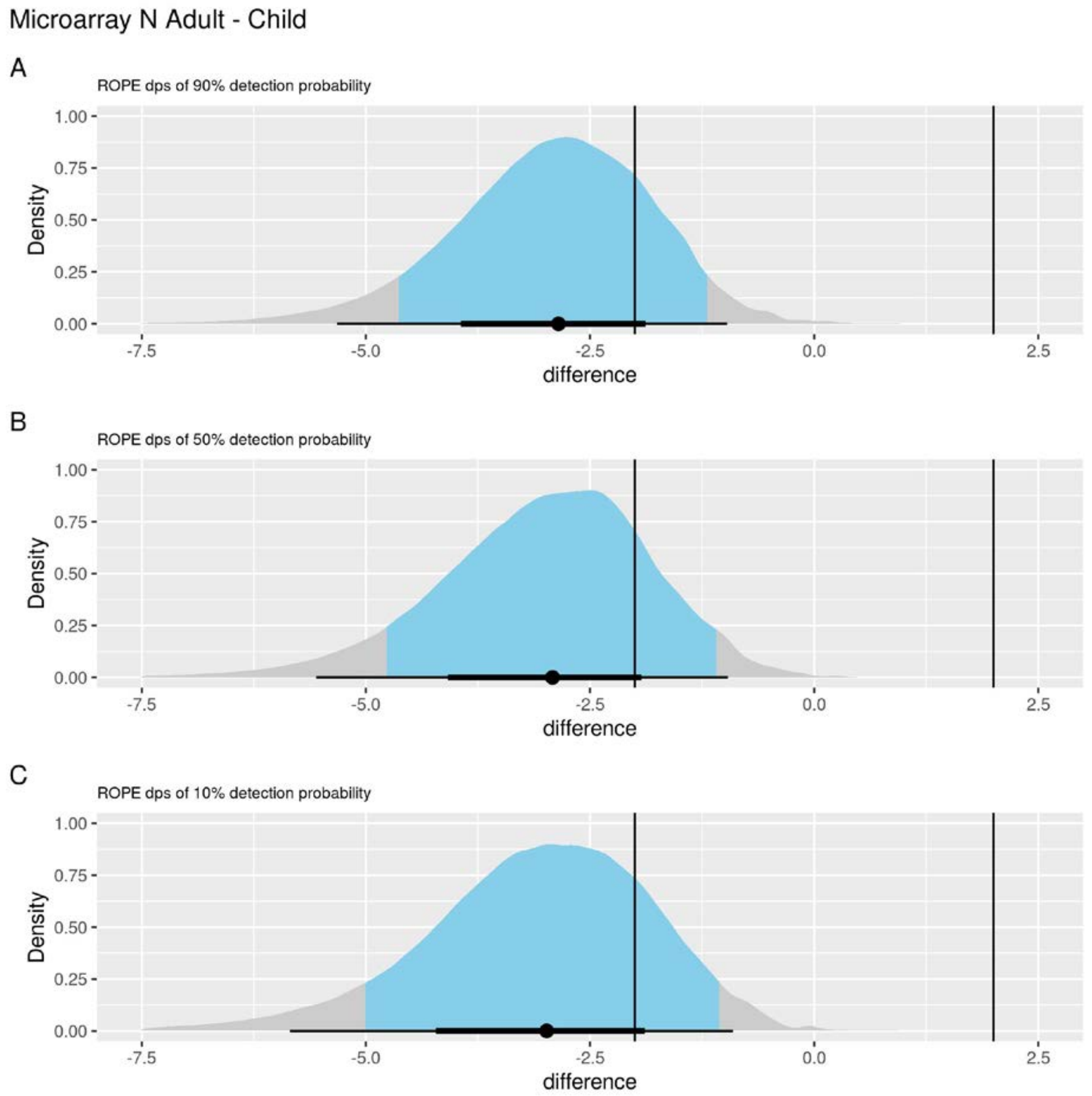


Figure S8: ROPE analyses for assessing differences between factors in Figure 6C and D. For serology detection probability (in dps) our rope interval is [-2,2], which means that we consider differences between days of less than 2 as not meaningful.

*Figure S9*

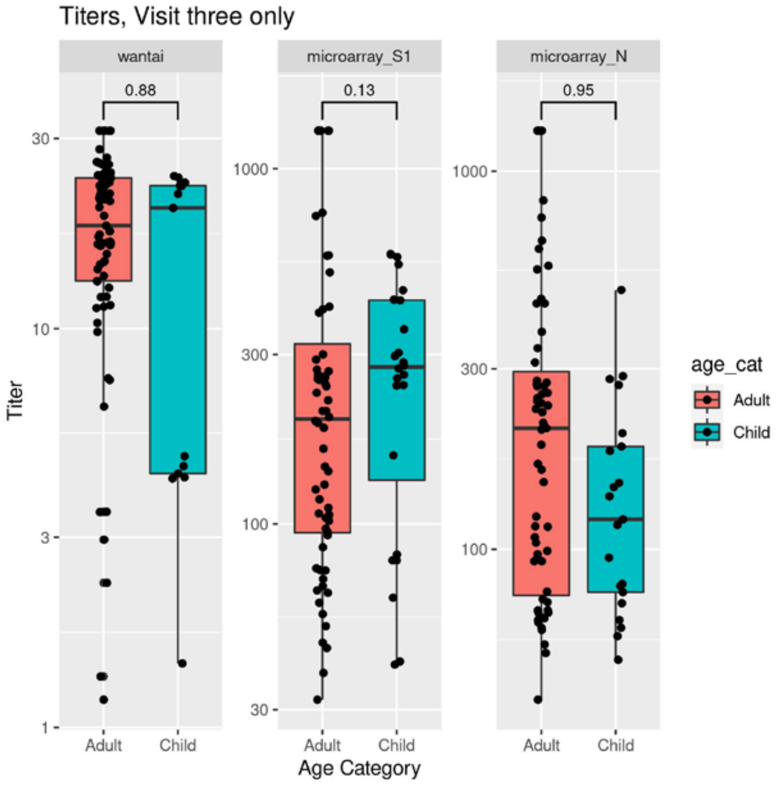


Figure S9: Effect of age on the development/magnitude of SARS-CoV-2 specific antibody responses. SARS-CoV-2 specific antibody titers at visit 3 (convalescence phase) in SARS-CoV-2 laboratory confirmed participants. SARS-CoV-2 Spike-specific IgM and IgG (Wantai, left), Spike-specific IgG (microarray S1, middle) and Nucleoprotein-specific IgG (microarray N, right) responses in relation to to age (adult and child). On the Y-axis, titers in OD ratio (Wantai) and EC50 values (microarrays) are indicated. The boxplot compactly displays the distribution of a continuous variable. It visualises five summary statistics (the median, two hinges and two whiskers). The lower and upper hinges correspond to the first and third quartiles (the 25th and 75th percentiles). The upper whisker extends from the hinge to the largest value no further than 1.5 * IQR from the hinge (where IQR is the inter-quartile range, or distance between the first and third quartiles). The lower whisker extends from the hinge to the smallest value at most 1.5 * IQR of the hinge. Differences were analyzed with the Wilcoxon test.
